# Search interest in alleged COVID-19 treatments during the pandemic and the impact of mass news media

**DOI:** 10.1101/2024.11.20.24317650

**Authors:** Emily E Ricotta, Samantha Bents, Brendan Lawler, Thomas Berkane, Fausto A Bustos Carrillo, Brianna A Smith, Maimuna S Majumder

## Abstract

**Background:** During a public health emergency, interest in unsafe or illegitimate medications can delay appropriate treatment and foster medical mistrust.

**Methods:** We obtained daily US-based Google Search Trends and Media Cloud data from 2019-2022 to assess the relationship between search interest and media coverage in three purported COVID-19 treatments: hydroxychloroquine, ivermectin, and remdesivir.

**Results:** Search interest and media coverage of all COVID-19 treatments were significantly elevated during the study period; search interest was highest for ivermectin (6.0 out of 100; interquartile range [IQR]: 1.9-9.9), while media covered hydroxychloroquine most frequently (0.05% of all articles published; IQR: 0.02-0.13%). Anomaly detection of both data sources identified several points of higher-than-expected activity; anomalies in search interest and media coverage showed similar patterns within treatments. There were distinct patterns of media coverage – while the plurality of sources for all treatments were considered “Left” or “Left Leaning”, ivermectin was covered by the highest number of “Right”-biased sources and remdesivir had the highest coverage by “Center” or “unbiased” sources. When assessing the co-occurrence of words and phrases in media sources covering each of the treatments, there were distinct qualitative difference in the categories of words appearing alongside the drugs. Specifically, ivermectin appeared to be reported more frequently in association with specific individuals than media mentioning the other drugs. In google searches, people seemed most interested in understanding what hydroxychloroquine is and the uses of ivermectin (e.g., “for humans”, “for dogs”). We found significant associations between media coverage and search interest for all three treatments. Media coverage had the strongest impact on same-day search interest for remdesivir (199.7% increase, 95% CI: 179.2, 221.6) and hydroxychloroquine (182.6% increase, 95% CI: 172.8, 192.7); interest dropped significantly 1 and 2 days after media coverage of these treatments. Interest in ivermectin was lower overall (105.0% increase, 95% CI: 97.9, 112.3) but stayed elevated even 2 days after media coverage. When evaluating the separate impact of search interest on media coverage, the associations were much weaker for all three treatments and all lagged conditions.

**Conclusions:** During a public health emergency, the information that populations access can directly influence health-seeking behaviors, with potentially life-threatening consequences. More broadly, positive media coverage of unsafe or unapproved medications can deter individuals from trusting and accessing safe alternatives that are more likely to be efficacious in preventing disease progression. Given the strong association between treatment-related news media coverage and public interest in said treatments, our results suggest that news media serve as a powerful mechanism for experts to inform the landscape of public opinion and to reach audiences during future public health emergencies.

## Introduction

Throughout the early COVID-19 pandemic, several repurposed drugs were evaluated as potential COVID-19 treatments in the United States, including hydroxychloroquine, ivermectin, and remdesivir. Hydroxychloroquine, an antimalarial and anti-rheumatic drug, gained US Food and Drug Administration (FDA) emergency use authorization to treat severe COVID-19 hospitalizations in March 2020; this was shortly revoked after clinical trials showed harmful cardiovascular side effects at the doses being dispensed.[1] Ivermectin, an antihelminthic, was proposed as an antiviral agent for COVID-19 in early 2020, leading to the initiation of several randomized controlled trials between mid-2020 and 2021;[2] however, results were generally inconsistent and inconclusive, leading the FDA, US National Institutes of Health, and World Health Organization to recommend against its use in 2021.[3] Remdesivir, an antiviral originally developed to treat hepatitis C, was approved by the FDA for persons hospitalized with COVID-19 in October 2020 after multiple clinical trials showed moderately improved outcomes among hospitalized cases.[4] Public interest in these treatments varied over time, driven partially by governmental, political, and media response to current events.[5] For hydroxychloroquine and then ivermectin, public demand for the drugs caused international pharmacy shortages, and in the case of ivermectin – a drug commonly used for livestock deworming – shortages of veterinary ivermectin as well.[6, 7] Beyond causing shortages, public interest in unsafe or untested/off-label medications during a public health emergency can delay access to appropriate treatments and foster mistrust in the medical system, a detriment at both individual and population levels. Therefore, both for situational awareness and interventional purposes, it is critical for health and policy communities to understand how the public obtains medical information during an emerging crisis.

One method of evaluating public interest in medical information is the use of internet search data. These data can serve as a large-scale, near real-time proxy for public interest by providing insight into what information people seek online, including the subject, timing, and frequency of specific topics. A recent study in Switzerland found that 55% of Google internet searches were informational (as opposed to transactional or navigational),[8] and in 2019, roughly 7% of Google searches (about 1 billion searches daily) were related to health.[9] This makes internet search data – publicly available through Google Search Trends (GST) – an attractive tool for health information research;[10] indeed, GST was used extensively throughout the pandemic to measure population engagement with public health messaging on topics like masking, vaccines, and misinformation, as well as interest in COVID-19 more broadly.[11, 12]

Another way to gauge public interest in a topic is by assessing relevant news media coverage – a data source that is particularly useful for topics like uncommon medications, which would rarely enter the public zeitgeist otherwise. The impact of news media on public interest and changing health behaviors is well studied,[13, 14] including during the COVID-19 era.[7, 11, 15] Previous research by our team and others have assessed a variety of health-related topics and their relationship with news media coverage using services like Media Cloud (MC),[16] an open-source content analysis tool that collects time series of national and sub-national news media coverage, comprising data from more than 50,000 news sources since its inception. The bulk of MC’s corpora are news stories from web-based media sites, which is particularly relevant as 86% of US adults reported getting news from an online source in 2023.[17]

Interestingly, the relationship between public interest and media coverage is not unidirectional; studies on communication theory demonstrate that the media can also serve as a reflection of public opinion rather than function solely as its driver – a phenomenon known as the “reflection hypothesis”.[18] As interest in a topic increases, people begin searching for things they see on the news, while in response, the news covers things they know people are interested in.[19]

Our study investigated the possibility of bidirectional relationships between online search interest and media coverage to evaluate exposure to and access of health-related information during the COVID-19 pandemic. Specifically, we examined: 1) the extent to which US news sources covered supposed COVID-19 treatments, 2) the extent of public interest in these treatments, as reflected by online search interest, and 3) the relationship between these data sources within the US.

## Methods

### Data sources for news media and internet searches

We used two primary data sources to assess the relationship between treatment-related news media coverage and treatment-related search interest in the US: MC and GST. All data were obtained for the period from January 2019 through December 2022 using the queries “hydroxychloroquine”, “ivermectin”, and “remdesivir” for both MC and GST. Data from 2019 were included in the analysis to provide a pre-pandemic baseline for each of these search terms; data collection ceased in 2022 after a sustained period of low search interest and media coverage of all three terms.

Daily treatment-related news media coverage data were downloaded from MC’s national- and state-level news corpora. These corpora include media articles published in either national (e.g., Reuters) or state-specific news sources (e.g., Arkansas Times) and report daily normalized content percentage (i.e., the number of corpus-specific articles reporting on a topic of interest on a given day divided by the total number of corpus-specific articles on the same day.)

Daily treatment-specific Google search interest data were downloaded using the Google Trends dashboard at the national and state level. Notably, search interest data are normalized by Google; the number of topic-specific searches is divided by the total searches conducted within a specified geography and time period, making GST data both fractional and relative to the region and time window selected. The data are scaled from 0-100, with a score of 100 representing peak search interest in the selected geography (i.e., state) and time interval. We obtained national-level GST data for anomaly detection and state-level GST for regression analysis, described below.

### National-level anomaly detection in treatment-specific search interest and media coverage

To identify peaks in treatment-related search interest and media coverage, we performed anomaly detection for each treatment in the respective time series at the national level using the a*nomalize* package in R,[20] with alpha set at 0.001, using the traditional seasonal decomposition by LOESS method for time series decomposition and the generalized extreme studentized deviate test for anomaly detection. For each detected anomaly, we conducted additional internet searches for potentially relevant current events that could have resulted in atypical media attention or public awareness.

In addition to strict media mentions, MC provides a list of top words in headlines of a random sample of content matching the query. These top words were used to create word clouds using the wordcloud2 R package after excluding the words “covid”, “covid-19”, and “coronavirus”.[21] Finally, we acquired bias ratings for the top 10 media sources for each treatment query primarily from Allsides.com;[22, 23] bias ratings were obtained from mediabiasfactcheck.com when Allsides.com did not rate a particular media source.[24] Ratings were pulled from mid-pandemic (2021) assessments when possible, or the closest assessment to that time when not.

### State-level association of treatment-specific search interest and media coverage

To visualize geographic heterogeneity in search interest and media coverage, we created heatmaps at the state level for each treatment using *ggplot2* (Supplemental Figure 1). Using treatment-specific regression models, we assessed the state-level relationship between treatment-specific media coverage and search interest. We considered three confounders in the models: political leaning, rurality, and social vulnerability.

### Covariates

As COVID-19 treatments were highly politicized during the initial stages of the pandemic,[9] we hypothesized that statewide political leaning may have differentially influenced interest in and coverage of specific treatments. We measured statewide political leaning as the percentage of a given state that voted for the Republican Party candidate in the 2020 presidential election, on a 0-100 scale.[25]

Social vulnerability may have also played a role in treatment interest given its association with reduced access to healthcare services.[26] Reduced interaction with healthcare providers and services has previously been associated with riskier health choices and poor health outcomes, providing evidence that social vulnerability may influence treatment-seeking behavior.[26, 27] We measured social vulnerability using the COVID-19 Community Vulnerability Index (CCVI), which provides a state-level measure of access to healthcare, taking economic, structural, and demographic barriers into account.[28] State-level scores are scaled continuously from 0-100, where 100 represents the highest level of vulnerability.

Lastly, we included rurality in the models, first as a proxy for known barriers to healthcare that could potentially influence off-label medication use,[29] and second because of ivermectin’s widespread use on farms as a livestock deworming agent, which we hypothesized could influence search interest especially among farming communities. Rurality was measured using the 2021 US Census Bureau Urban and Rural dataset, which provides the percentage of a given state’s population that lives in rural settings, measured on a 0-100 scale, with 100 being the most rural.[30]

Additional covariates were considered for inclusion in this analysis and were ultimately rejected. Specifically, we did not feel that broadband (BB) access is a confounding factor in the relationship between GST and MC as BB access is unlikely to influence whether the media covers COVID-19 treatments (Supplemental figure 2). However, for due diligence, we did acquire a BB access database (US FCC Broadband Data Collection National Broadband Map)[31, 32] for May 2024 (the earliest version of the dataset we could locate) and discovered that the median percentage of the US served by BB (≥100/20 Mbps) is 94%, the median percentage for underserved (≥25/3 Mbps but <100/20 Mbps is 5%, and the median unserved (<25/3 Mbps) is 0%. These numbers were similar when stratifying by urban/rural. We therefore chose not to include BB access in our analysis.

### Analysis

Fitting zero-inflated negative binomial models to account for overdispersion in both variables when serving as the exposure, we regressed search interest against weekly media coverage, and vice-versa, for each treatment using the *glmmTMB* package in R.[33] Zero-inflated models were chosen because across most days and states in the dataset outside of the anomalies, there was little search interest or media coverage of the selected COVID-19 treatments. As a sensitivity analysis, we attempted to refit the data using beta regression models (accounting for continuous rather than count outcomes). However, GST data are pre-scaled to the [0, 1] interval by Google, and beta models in *glmmTMB* can only accommodate outcomes on the [0, 1) interval. It was therefore not possible to incorporate beta models into our analytic approach.

Media coverage was scaled by dividing the value by the root mean square, and the other covariates were mean-centered and scaled by dividing the value by the standard deviation to ensure comparability and equal contribution in the regression analysis and aide in interpretation. We included “state” as a random intercept to control for interstate variability in the impact of covariates on the exposure. Media coverage and search interest were assessed at three relative time points: non-lagged (same day), lagged one day, and lagged two days using the *timetk* R package.[34] Multicollinearity was assessed by checking the variance inflation factor of the models. Model results are presented as percent difference in search interest with 95% Wald confidence intervals. Multiple comparisons were accounted for by using the Bonferroni method, with p < 0.002 considered significant. All analyses were performed using R version 4.4.x.[35]

### Data availability

All data used in this study are available through public databases. Project-specific GST and MC data along with annotated study code can be found on Github.[36]

## Results

### National trends in search interest and media coverage

Nationally, the median search interest (out of 100) during the pandemic period (January 2020–December 2022) was 2.2 for hydroxychloroquine (interquartile range [IQR]: 1.6-3.0), 6.0 for ivermectin (IQR: 1.9-9.9), and 2.2 for remdesivir (IQR: 1.5-3.3), compared to 0.5 (IQR: 0.3-0.6), 1.0 (IQR: 1-1), and 0 (IQR: 0-0) for each treatment, respectively, in 2019. Nationally, the median media coverage during the pandemic was 0.05% for hydroxychloroquine (IQR: 0.02-0.13%), 0.01% for ivermectin (IQR: 0-0.04%), and 0.03% for remdesivir (IQR: 0.01-0.1%), compared to 0 median coverage for each of the treatments in 2019.

Anomaly detection identified periods of atypical search interest (Figure 1a) and media coverage (Figure 1b). Search interest in hydroxychloroquine peaked in July 2020 with media coverage of it increasing sharply in March 2020 and remaining high until mid-November 2020. While search interest in ivermectin had several small anomalies prior to 2021, interest peaked in late August/early September 2021, with a second large peak occurring in January 2022. Media coverage of ivermectin peaked and stayed elevated during this same period. Finally, search interest in remdesivir peaked in late April 2020 and decreased quickly; media coverage was high throughout 2020, and coverage remained sporadically elevated through the end of the study period.

**Figure 1.**
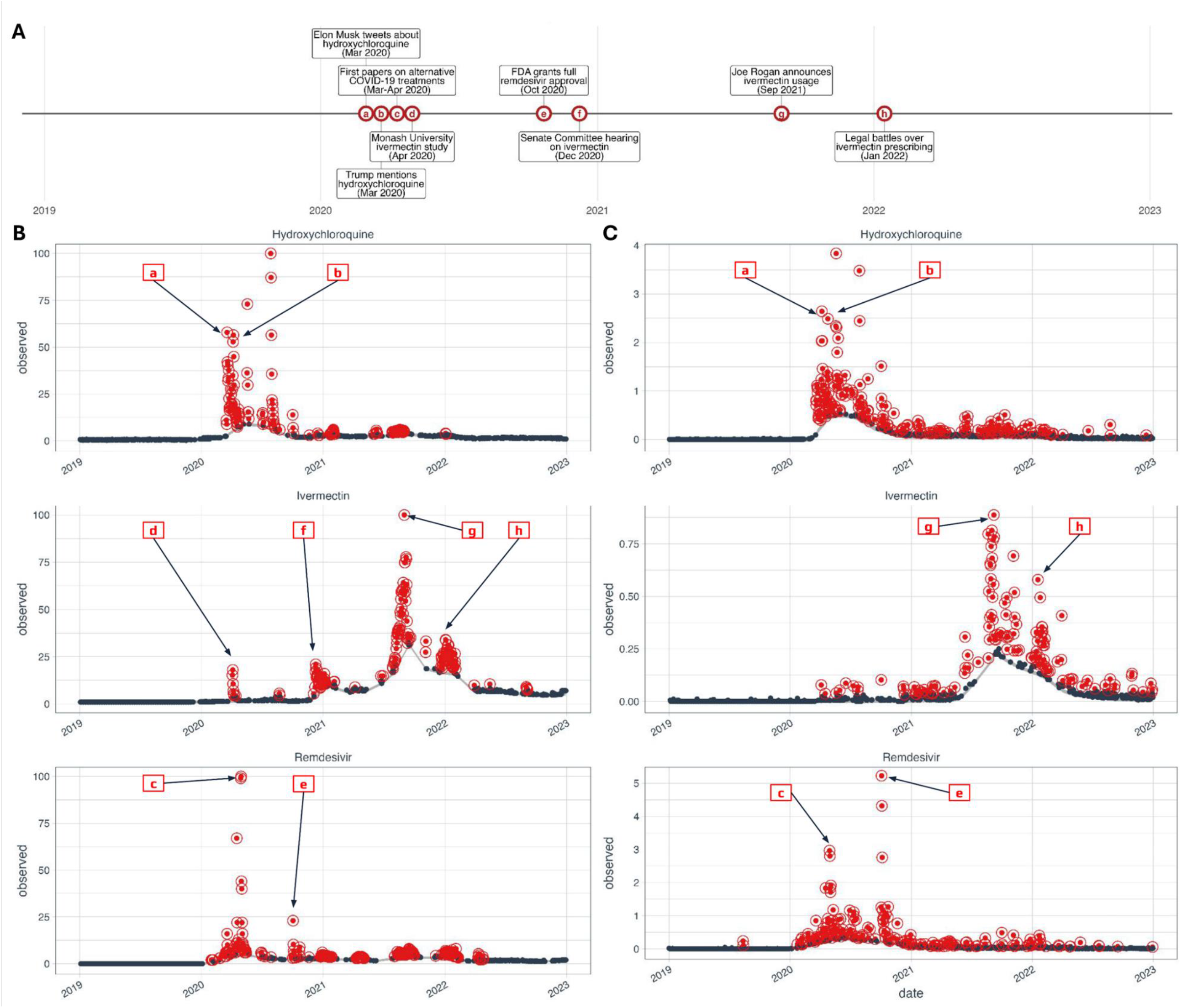
National GST and MC time series for hydroxychloroquine, ivermectin, and remdesivir from 2019-2022. (A) Timeline of potentially relevant current events. Identified anomalies are shown in red and non-anomaly time points are shown in gray for (B) GST and (C) MC. Annotated with COVID-19 treatment-related events occurring contemporaneously with the anomalies. Note that the y-axis differs across treatments in column B, in contrast to column A, which is always scaled from 0 to 100 by Google. Annotations were located by web searching for relevant news events contemporaneous to detected anomalies. Abbreviations: GST = Google Search Trends, MC = Media Cloud.

Table 1 shows the top 10 national-level media sources with the highest number of stories matching each treatment query and their bias as determined by one of two media bias reporting websites during the COVID-19 pandemic. Unsurprisingly, all top sources were web-based. While some sources covered all treatments at a high level (e.g., nytimes.com and washingtonpost.com were in the top 10 for all three treatments), there were distinct patterns of bias (Figure 2). The plurality of sources for all treatments were considered “Left” or “Left Leaning”; ivermectin was covered by the highest number of “Right”-biased sources, and remdesivir had the highest coverage by “Center” or “unbiased” sources.

**Table 1.** Top 10 media sources for each treatment query during the study period (2019-2022). Each source is listed with the number of stories mentioning the treatment. The political bias of each source and the year of assessment were obtained from AllSides or Media Bias/Fact Check.[22–24]

| Hydroxychloroquine |  |  |  | Ivermectin |  |  |  | Remdesivir |  |  |  |
| --- | --- | --- | --- | --- | --- | --- | --- | --- | --- | --- | --- |
| Source | Count | Bias | Year | Source | Count | Bias | Year | Source | Count | Bias | Year |
| reuters.com | 1498 | C | 2021 | nytimes.com | 357 | LL | 2021 | reuters.com | 1777 | C | 2021 |
| cnn.com | 1402 | L | 2021 | washingtonpost.com | 190 | LL | 2020 | nytimes.com | 1049 | LL | 2021 |
| nytimes.com | 1042 | LL | 2021 | rawstory.com* | 173 | L | 2025 | cnn.com | 889 | L | 2021 |
| washingtonpost.com | 935 | LL | 2020 | theguardian.com | 170 | LL | 2021 | cbsnews.com | 468 | LL | 2021 |
| rawstory.com* | 628 | L | 2025 | newsweek.com | 154 | C | 2021 | cnbc.com | 450 | C | 2022 |
| foxnews.com | 600 | R | 2021 | foxnews.com | 145 | R | 2021 | marketwatch.com | 429 | C | 2020 |
| theguardian.com | 548 | LL | 2021 | cnn.com | 142 | L | 2021 | washingtonpost.com | 422 | LL | 2020 |
| usatoday.com | 460 | LL | 2021 | dailykos.com* | 139 | L | 2024 | thestreet.com* | 397 | RL | 2020 |
| cbsnews.com | 379 | LL | 2021 | theblaze.com | 129 | R | 2023 | businessinsider.com | 362 | LL | 2022 |
| businessinsider.com | 365 | LL | 2022 | forbes.com | 119 | C | 2022 | benzinga.com* | 360 | C | 2021 |
| *Rating from Media Bias/Fact Check Abbreviations: C = Center, L = Left, LL = Left Leaning, R = Right, RL = Right Leaning |  |  |  |  |  |  |  |  |  |  |  |

**Figure 2.**
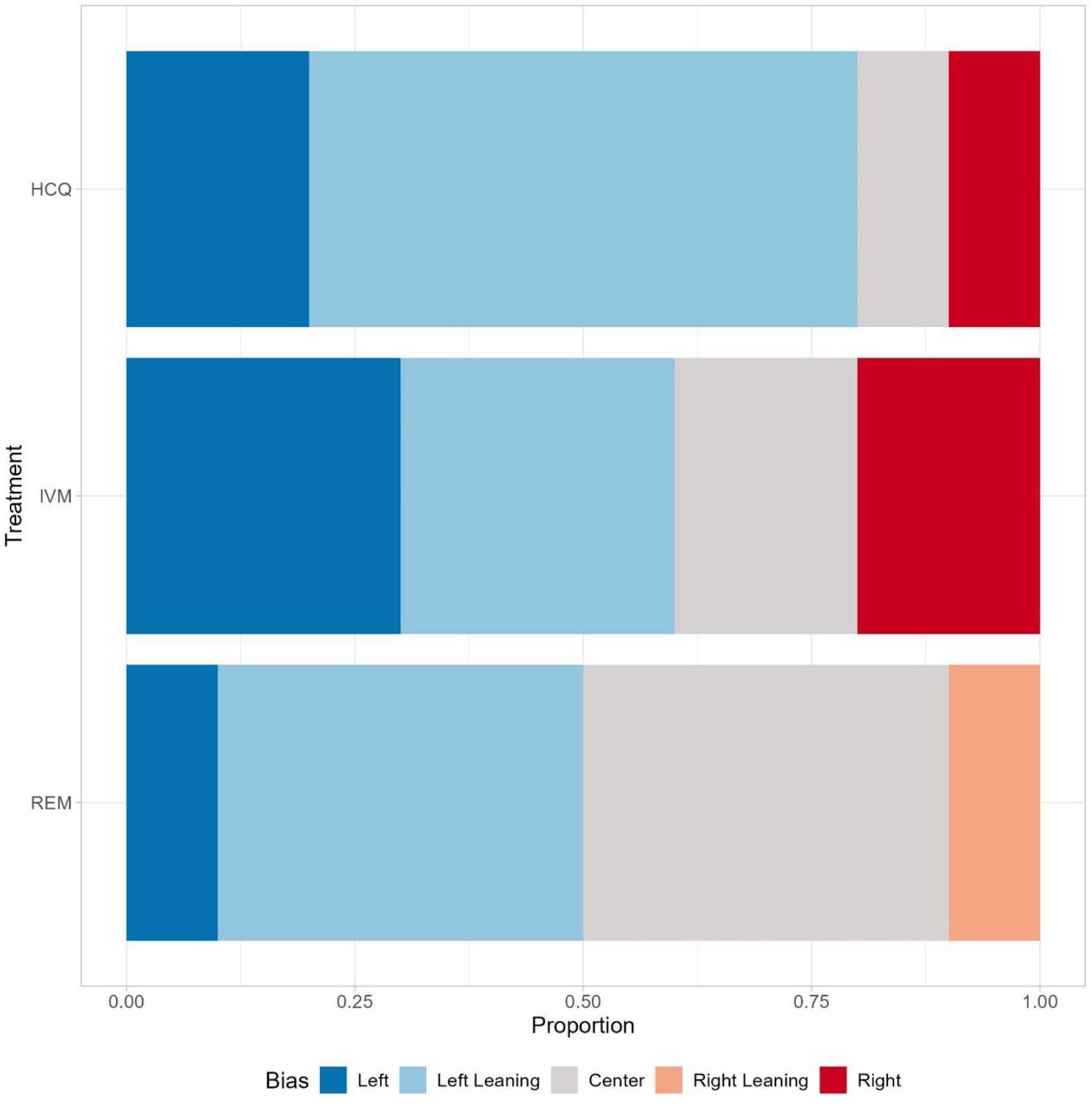
Bias category for top 10 media sources covering each purported COVID-19 treatment during the pandemic period (2019-2022). The political bias of each source and the year of assessment were obtained from AllSides or Media Bias/Fact Check.[22–24] Abbreviations: HCQ = hydroxychloroquine, IVM = ivermectin, REM = remdesivir.

When assessing the co-occurrence of words and phrases in media sources covering each of the treatments (Figure 3a-c), the most striking qualitative difference was in the categories of words appearing alongside the drugs. Specifically, ivermectin appeared to be reported more frequently in association with specific individuals (Figure 3b) than media mentioning hydroxychloroquine (Figure 3a) or remdesivir (Figure 3c), although the top co-occurring word for hydroxychloroquine was “Trump” by a wide margin. In google searches, people seemed most interested in understanding what hydroxychloroquine is and the uses of ivermectin (e.g., “for humans”, “for dogs”). Curiously for remdesivir, several co-occurring terms included the manufacturer and searches for remdesivir stock, rather than its uses (Figure 3d).

**Figure 3.**
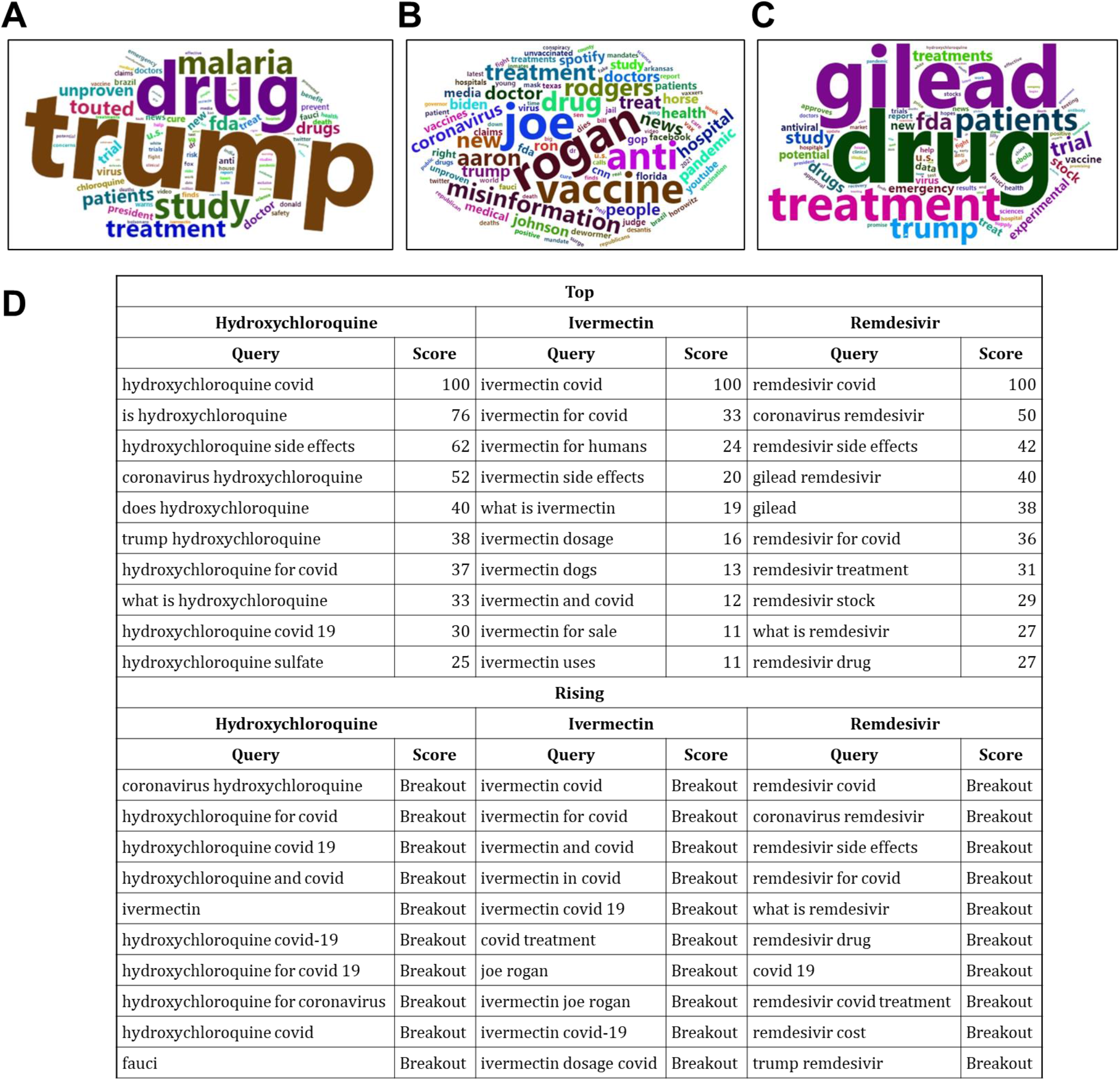
Co-occurrence of words and phrases in media sources (A-C) and GST (D) covering each of the treatments. (A-C) Word clouds featuring the top words in headlines of a random sample of content matching the query (i.e., “hydroxychloroquine”). (D) Table of co-occurring search terms. Per Google: “Top searches are terms that are most frequently searched with the term you entered in the same search session, within the chosen category, country, or region. Rising searches are terms that were searched for with the keyword you entered, which had the most significant growth in volume in the requested time period. For each rising search term, you see a percentage of the term’s growth compared to the previous time period. If you see “Breakout” instead of a percentage, it means that the search term grew by more than 5000%.”

### State-level trends in search interest and media coverage

When controlling for average state political leaning, rurality, and social vulnerability, we found significant associations between media coverage and search interest for all three treatments (Figure 4). Media coverage had the strongest impact on same-day search interest for remdesivir (199.7% increase, 95% CI: 179.2, 221.6) and hydroxychloroquine (182.6% increase, 95% CI: 172.8, 192.7); interest dropped significantly 1 and 2 days after media coverage of these treatments. Interest in ivermectin was lower overall (105.0% increase, 95% CI: 97.9, 112.3) but stayed elevated even 2 days after media coverage (Figure 4a). Interestingly, increasing CCVI was associated with increased search interest in all three treatments, sustained across the time lag (Figure 4c). State rurality was negatively associated with search interest for ivermectin across all time lags, and state political leaning was negatively associated with interest in remdesivir (Figure 4c). When evaluating the separate impact of search interest on media coverage, the associations were much weaker for all three treatments and all lagged conditions. Same-day search interest for remdesivir had the strongest impact on media coverage, with one standard deviation increase in same-day search interest being associated with an 18.1% increase (95% CI: 17.0, 19.2) on media coverage. The comparable effect sizes for hydroxychloroquine and ivermectin were significantly lower at 12.4% and 8.7%, respectively.

**Figure 4.**
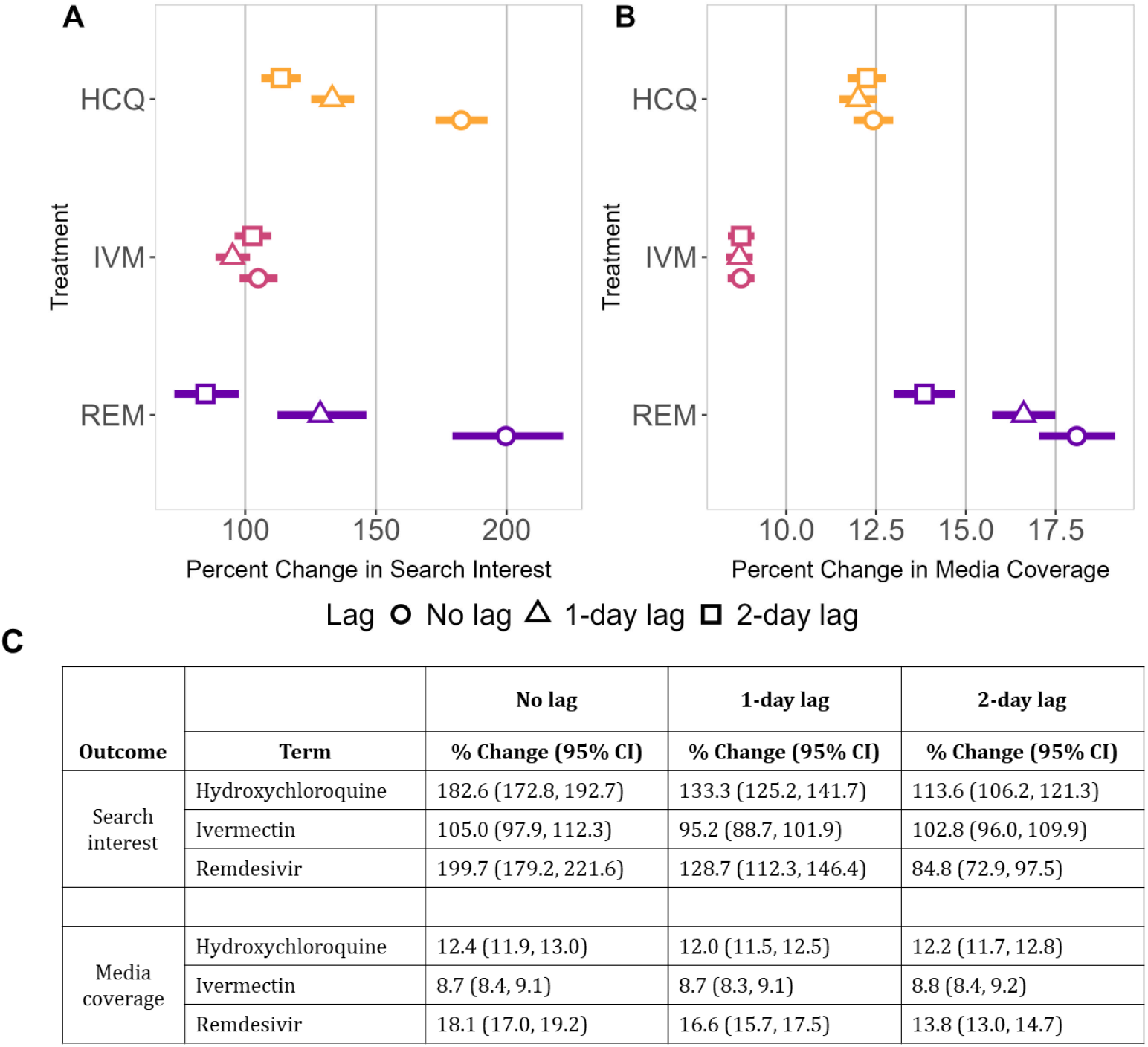
Output from same-day, 1-day, and 2-day lagged regressions, looking at the effect of (A) media coverage on search interest and (B) search interest on media coverage. (C) presents the results in tabular format as the percent change in outcome for one standard deviation change in the exposure with the associated 95% confidence intervals. Abbreviations: HCQ = hydroxychloroquine, IVM = ivermectin, REM = remdesivir.

## Discussion

In this study we identified significant public interest in three purported COVID-19 treatments – hydroxychloroquine, ivermectin, and remdesivir – as represented by anomalies detected in heightened search interest these medications during the pandemic period. Additionally, we found significant associations between news media coverage and all three treatments. Importantly, we observed that media coverage was a stronger driver of search interest than search interest was of media coverage.

At the beginning of the pandemic, substantial uncertainty surrounded the epidemiology of COVID-19 and the effectiveness, safety, and availability of potential treatments. As scientific evidence accumulated, public health guidance evolved accordingly to reflect updated knowledge.[37] Although changing recommendations are expected during the early stages of a novel pathogen outbreak, inconsistent or insufficiently transparent messaging may have contributed to public confusion. In this environment, many individuals increasingly relied on non-expert sources, such as politicians, public figures, and celebrities, for health-related information rather than traditional public health authorities.[5] Scholars have long noted that media reporting practices can further shape public risk perception; anecdotal framing and selective emphasis may lead audiences to draw inaccurate conclusions about the magnitude or nature of risk.[38] Additionally, because many journalists lack formal training in statistics or probability, complex or preliminary scientific findings may not always be critically evaluated before dissemination to mass audiences, potentially amplifying misunderstanding or misinterpretation.[39]

In addition, recent studies report that the percentage of Americans who regularly see a primary care physician has steadily decreased between 2014 and 2022, meaning regular communication with licensed medical providers is becoming more limited for many communities across the US.[40] These factors may have contributed to the general public’s increasing reliance on other information sources – such as news media – for guidance during the COVID-19 pandemic. Indeed, over the last several years, perceived US media influence on government decisions has grown among the public and will likely continue to have an outsized influence on public interest.[41] In communication research, this aligns with the media system dependency theory, which assumes that the impact of information provided by news media on audience perceptions is a function of how dependent an audience member is on mass media as a source of acquiring information that is important to them, specifically;[42] that is, the more likely someone is to turn to news media to obtain personally relevant information, the more *dependent* on the media as a source they become. Importantly, research has found that this dependency on media for information is what drives changes in behavior, not media usage alone. While our study did not directly assess media usage or dependency at the individual level, the patterns observed are consistent with media system dependency theory and highlight the importance of integrating communication theory into future epidemiologic analyses of health information seeking.

We found that the effect of media coverage significantly increased same-day search interest for all three treatments. This effect was largest for remdesivir, the only treatment that remains FDA-approved for use,[3, 4] and hydroxychloroquine, an initially FDA-authorized treatment with early potential that ultimately proved ineffective and dangerous.[1] While the effect was overall more moderate for ivermectin, an incredibly popular but inappropriate treatment for COVID-19, it was the only drug for whom search interest stayed significantly elevated to the same level one to two days after coverage by media outlets. One possible explanation for our findings is that people may receive information about ivermectin more frequently through non-news media sources, such as personal networks, social media, or podcasts, which may see continued high levels of contact engagement days to weeks following posting, depending on the platform.[43] Another hypothesis for why the impact of media coverage on search interest for ivermectin was low compared to the other therapies relates to trust in traditional media. Research by Ladd (2012) suggests that individuals who distrust traditional news media sources are more likely to rely on preexisting political beliefs to form opinions, rather than incorporating new information supplied by news media.[44] At least one study has found that higher belief in COVID-19 vaccine misinformation and conspiracy thinking was associated with increased use of ivermectin and/or hydroxychloroquine,[45] although the results were unfortunately not stratified by specific treatment. In a more recent article by Ladd and Podkul (2020), they argue that when media credibility is lost, so is its capacity to influence the public.[46] In this context, diminished trust in traditional media and reliance on alternative information channels may attenuate or delay the observable impact of news coverage on public search behavior.

We note several limitations in this analysis. First, the state-level analysis includes only state-level media outlets, as MC cannot track state-level readership of national news sources. Given that many individuals rely heavily on national media outlets for news, the media exposure used in this analysis may be skewed if individuals within a state engage differently with national-versus state-level news outlets. Additionally, larger states had relatively larger news media corpora than smaller states; however, no corpus reported fewer than 30 sources, indicating that sample sizes were sufficient to assess news media coverage at this geographic scale. Notably, MC does not capture sentiment (i.e., the emotions and attitudes contextualizing a news report about a particular issue), meaning we were unable to comment on whether the treatment-related news media sources were reporting positively or negatively on purported treatments. Future studies should assess the content and sentiment of health-related news articles to understand the kind of language that is most likely to prompt heightened search interest so those techniques can be leveraged to improve health communication strategies. Lastly, public interest was only captured for the population using Google as their primary search engine; people using alternate engines like Bing or DuckDuckGo were not captured in our study and these users might differ in characteristics like political leaning that could make their search behavior fundamentally different from Google users. However, in the US, the Google search engine averages nearly 90% of the market share;[47] we are therefore confident that our results capture most relevant search interest.

## Conclusions

Regardless of the quality of public health messaging available to the general public, social media, news outlets, as well as political and public figures will continue to be major influences on how individuals seek health-related guidance. During a public health emergency, the information that populations access can directly influence health-seeking behaviors, with potentially life-threatening consequences. More broadly, positive media coverage of unsafe or unapproved medications can deter individuals from trusting and accessing safe alternatives that are more likely to be efficacious in preventing disease progression. Given the strong association between treatment-related news media coverage and public interest in said treatments, our results suggest that news media serve as a powerful mechanism for experts to inform the landscape of public opinion and to reach audiences during future public health emergencies.

## Data Availability

All data used in this study are available through public databases.

https://trends.google.com/trends/

https://www.mediacloud.org/

https://www.census.gov/data/tables/time-series/demo/voting-and-registration/p20-585.html

https://www.census.gov/programs-surveys/geography/guidance/geo-areas/urban-rural.html

https://covid-static-assets.s3.amazonaws.com/US-CCVI/COVID-19+Community+Vulnerability+Index+(CCVI)+Methodology.pdf

## Author Contributions

MSM and EER conceptualized this article. SB, BL, EER, and TB were responsible for data acquisition. SB, BL, and EER conducted data analysis and created all figures. All authors contributed to data interpretation. SB, BL, and EER wrote the initial draft of the manuscript. EER, BS, and FABC wrote the second draft. All authors contributed to its editing and final version.

## Funding

EER and SB were supported in part by the Division of Intramural Research, National Institute of Allergy and Infectious Diseases, National Institutes of Health. EER and FABC were supported by departmental startup funds from the Department of Preventive Medicine and Biostatistics, Uniformed Services University of the Health Sciences. BL was supported in part by grant SES2230083 from the National Science Foundation. MSM was supported in part by grants SES2200228 and IIS2229881 from the National Science Foundation. TB was supported in part by a Moderna Fellowship Award. The CompEpi Dispersed Volunteer Research Network is sponsored in part by grant R35GM146974 from the National Institute of General Medical Sciences, National Institutes of Health.

## Disclaimers and Conflict of Interest

The contents of this publication are the sole responsibility of the authors and do not necessarily reflect the views, opinions, or policies of the National Institutes of Health, the Uniformed Services University of the Health Sciences, the US Department of Health and Human Services, the US Department of War, the authors’ funders, or the US Government. This work was prepared by a civilian employee of the US Government as part of the individual’s official duties and therefore is in the public domain and does not possess copyright protection (public domain information may be freely distributed and copied; however, as a courtesy it is requested that the authors be given an appropriate acknowledgement). Authors declare no financial interest in any commercial product, service, or organization providing financial support for this research. References to non-Federal entities or products do not constitute or imply a Department of War or Uniformed Services University of the Health Sciences endorsement.

---

**Supplemental Figure 1.**
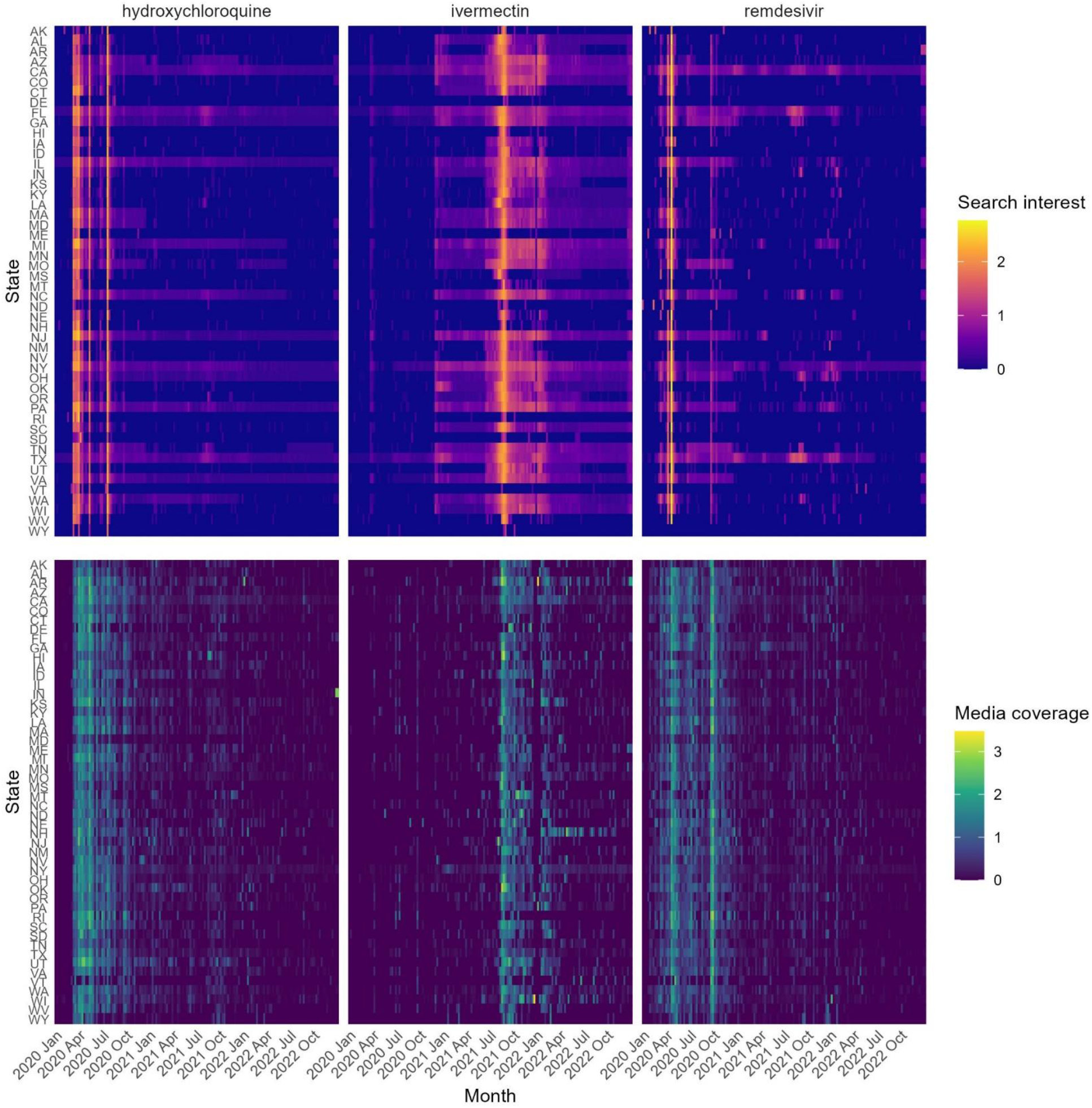
Search interest and media coverage by state for each of the three treatments. The weekly average search interest (top) and media coverage (bottom) were log transformed and plotted weekly from January 2020 through October 2022. Each row represents a state time series. Warmer colors represent a higher average.

**Supplemental Figure 2.**
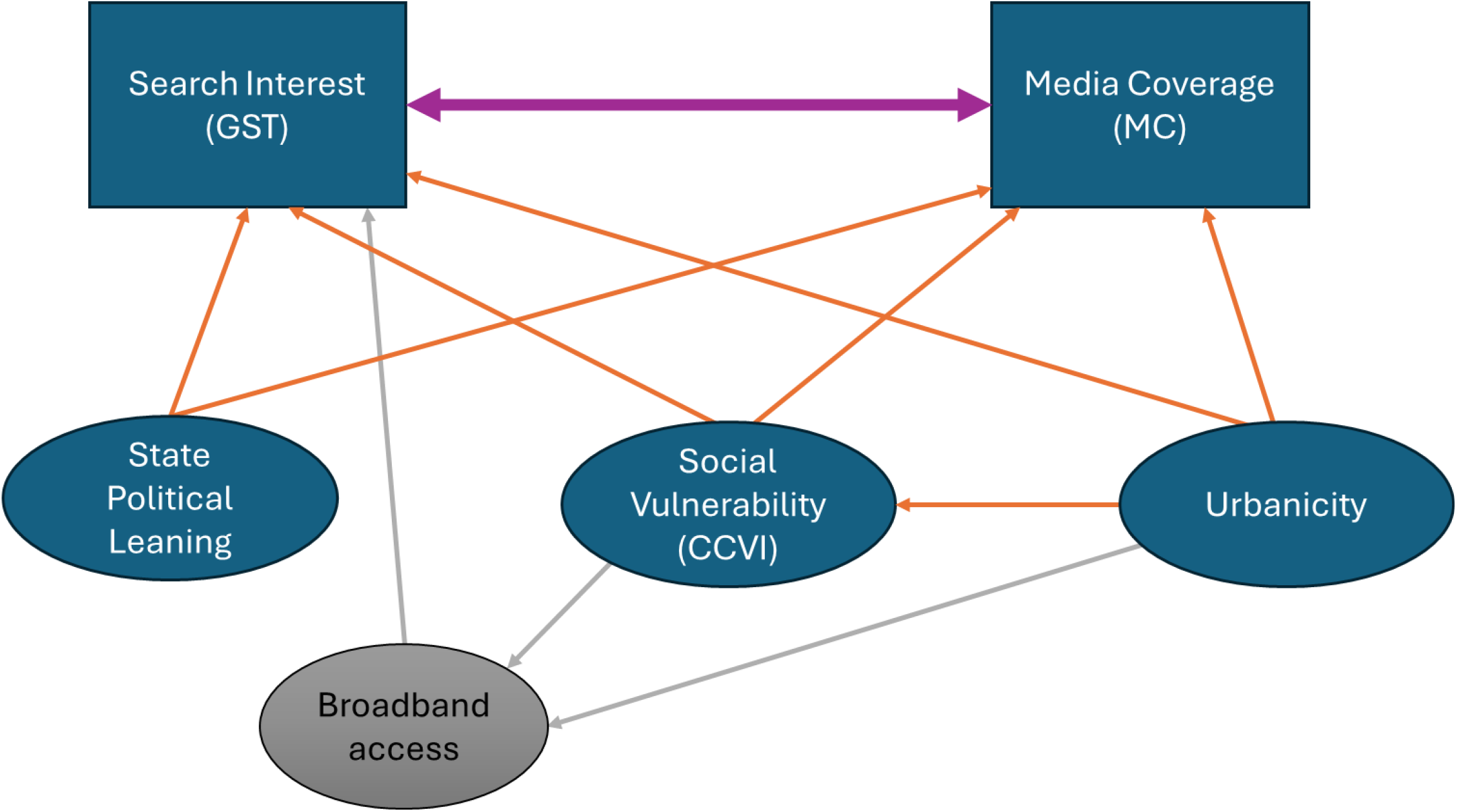
A directed acyclical graph depicting the hypothesized relationship between GST, MC, and several potential confounders. Ultimately, broadband access was ruled out as a confounder and thus not included in the final analysis. Abbreviations: CCVI = COVID-19 Community Vulnerability Index, GST = Google Search Trends, MC = Media Cloud.

## Notes

### Competing Interest Statement

The authors have declared no competing interest.

### Summary of Updates

This version of the manuscript has undergone substantial re-analysis, including new data and additional analytic methods.

