## Supplemental Figures for "Search interest in alleged COVID-19 treatments during the pandemic and the impact of mass news media"

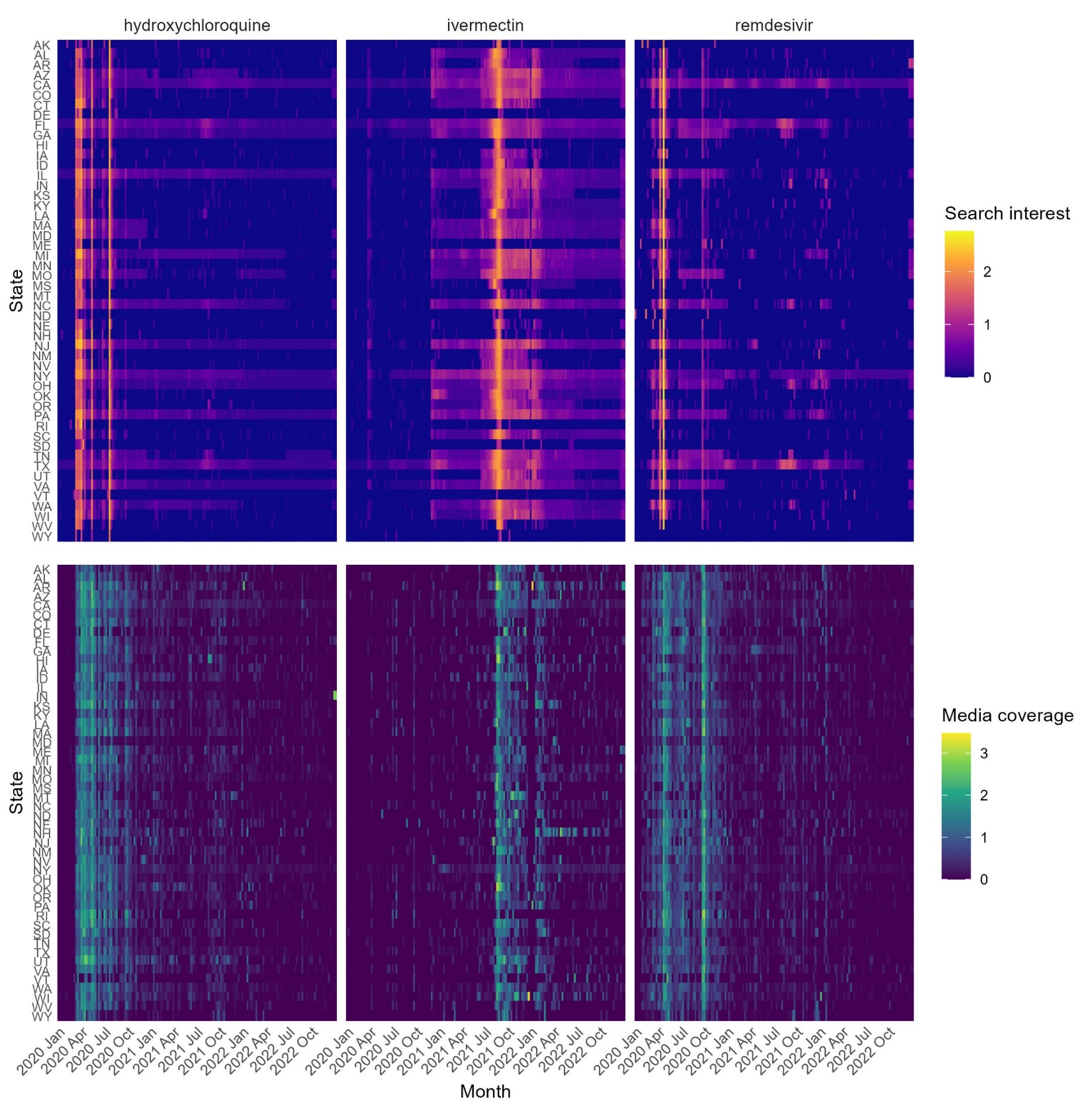

**Supplemental Figure 1. Search interest and media coverage by state for each of the three treatments.** The weekly average search interest (top) and media coverage (bottom) were log transformed and plotted weekly from January 2020 through October 2022. Each row represents a state time series. Warmer colors represent a higher average.

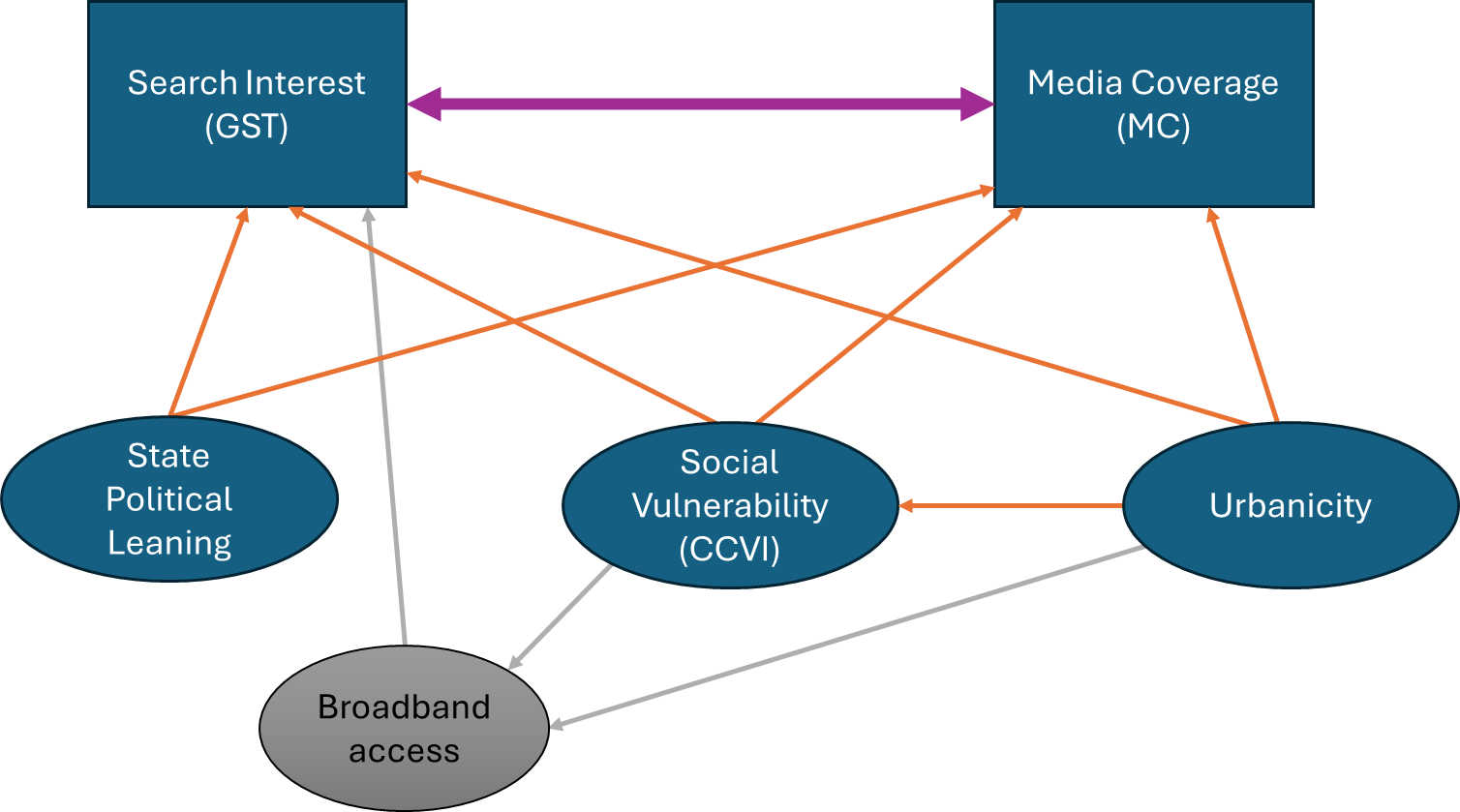

**Supplemental Figure 2. A directed acyclical graph depicting the hypothesized relationship between GST, MC, and several potential confounders.** Ultimately, broadband access was ruled out as a confounder and thus not included in the final analysis. Abbreviations: CCVI = COVID-19 Community Vulnerability Index, GST = Google Search Trends, MC = Media Cloud.
